# DOXYCYCLINE TO TREAT LEVODOPA-INDUCED DYSKINESIAS IN PARKINSON’S DISEASE: A PROOF-OF-CONCEPT STUDY

**DOI:** 10.1101/2022.05.13.22275023

**Authors:** Bruno Lopes Santos-Lobato, Manuelina Mariana Capellari Macruz Brito, Ângela Vieira Pimentel, Rômulo Torres Oliveira Cavalcanti, Elaine Del-Bel, Vitor Tumas

## Abstract

**Background:** Levodopa-induced dyskinesia (LID) is a common motor complication of levodopa therapy in patients with Parkinson’s disease (PD). Doxycycline is a widely used and inexpensive tetracycline with anti-inflammatory properties.

**Objective:** Evaluate the efficacy and safety of doxycycline in patients with PD and LID.

**Methods:** This was an open-label, single-center, phase 2 proof-of-concept study in patients with PD with mild functional impact of dyskinesia, which used levodopa three times daily, in a movement disorders clinic in Brazil. Participants were treated with doxycycline 200 mg/day for 12 weeks, with evaluations in baseline, week 4, and week 12 of treatment. The primary outcome measure was the change from baseline in the Unified Dyskinesia Rating Scale (UDysRS) total score at week 12, evaluated by two blinded raters. Key secondary outcomes measures were OFF time and ON time with troublesome dyskinesia in the PD home diary.

**Results:** Eight patients with PD were treated and evaluated. Doxycycline 200 mg/day reduced the UDysRS total score in week 12, compared with baseline (Friedman’s X^2^ = 9.6, p = 0.008). Further, doxycycline reduced the ON time with troublesome dyskinesia (Friedman’s X^2^ = 10.8, p = 0.004) without worsening parkinsonism. There were no severe adverse events, and dyspepsia was the commonest event.

**Conclusions:** Doxycycline was effective in reducing LID and safe after a 12-week treatment. Further well-designed placebo-controlled clinical trials with a longer duration and a larger number of participants are needed.

## INTRODUCTION

Parkinson’s disease (PD) is the second most common neurodegenerative disease, affecting approximately 6.1 million people worldwide^1^. Levodopa is the gold standard medical therapy for motor symptoms (bradykinesia, rigidity, tremor, gait disorders)^2^. However, chronic treatment with levodopa causes the onset of levodopa-induced dyskinesia (LID), hyperkinetic involuntary movements temporally associated with the use of levodopa^3^.

The pathophysiology of LID is still not completely understood, but recent evidence has shown that neuroinflammation might be a leading cause of the onset of this motor complication. Animal models of dyskinesia have shown that the onset of abnormal involuntary movement in rats is associated with an increase in the density of astrocytes and activated microglia in the striatum, as well as a higher expression of proinflammatory mediators (GFAP, OX-42, TNF-α, iNOS) in neurons and glia of dyskinetic animals^4,5^. In humans, there were high levels of a mediator of inflammation, the arachidonic acid, in the temporal cortex of postmortem brains of patients with PD who had developed motor complications (including LID), compared to healthy controls and patients with PD without motor complications^6^. Recent CSF analyses of patients with LID in PD revealed some neuroinflammatory abnormalities: a distinct metabolic profile strongly related to the dysregulation of lipid metabolism^7^ and high levels of nitrite and nitrate, which may be associated with the increased intrathecal production of nitric oxide by astrocytes and microglia^8^. These results suggest that a chronic pro-inflammatory state in the brain may be associated with LID onset^9^. Although it is unclear what drives this increased inflammation, targeting neuroinflammation might be a pharmacological strategy to limit this motor complication.

Until now, the only effective medication to reduce LID in PD without worsening parkinsonism is amantadine^10^, which may cause psychosis and livedo reticularis. Amantadine has been recently shown to possess anti-inflammatory effects^11^. Many clinical trials tested the efficacy of different drugs in LID, with inconclusive results^12^. As an alternative for this shortage of treatments for LID management, repurposing drugs with proven safety may be an efficient method to bring new therapies to patients^13^. Among the most investigated drugs for repurposing, antibiotics have been tested in neurodegenerative disease due to their several mechanisms of action through modulating signaling pathways^14^.

Doxycycline is an inexpensive second-generation semisynthetic tetracycline with easy penetration of the blood-brain barrier, reduced toxicity, longer half-lives, superior tissue fluid penetration^15^, with exceptional bioavailability^16^. Despite being commonly used as an antibiotic to treat infectious diseases, doxycycline is also prescribed as an anti-inflammatory drug in the management of acne vulgaris and rosacea. Indeed, the therapeutic rationale for targeting neuroinflammation is further supported by the observation of a reduced risk of PD in individuals using tetracyclines for rosacea treatment^17^. The treatment with doxycycline is associated with few adverse events, particularly gastrointestinal symptoms and skin reactions, even after long-term administration^18^. However, there are some concerns about its long-term treatment, such as antibiotic resistance and interference on gastrointestinal microbiota.

Doxycycline has an anti-inflammatory effect in the nervous system, based on inhibition of glial activation in the substantia nigra and the striatum and suppression of metalloproteinase induction^19^. Also, doxycycline reduces the transcription of proinflammatory mediators suppressing the p38 MAPK and NF-κB signaling pathways^15^, and restraining angiogenesis^20^.

Previously, doxycycline reduced the dopaminergic cell loss in rodent PD models^19^, and inhibited microglia activation in an in vitro model of neuroinflammation^15^. Regarding LID, a recent study showed that acute and chronic intraperitoneal administration of doxycycline reduced the onset of dyskinesias in a rat model of LID. Furthermore, doxycycline reduced the expression of molecular markers of LID (FosB, COX-2, GFAP, OX-42) in the striatum of rats that developed dyskinesias. A derivative of doxycycline without antibiotic properties, COL-3, also reduced the onset of dyskinesias^21^.

Considering the low cost and good safety of doxycycline and its modulating effect on neuroinflammatory mechanisms, it may represent a new therapy for LID management. Thus, we conducted an open-label, proof-of-concept phase 2 clinical trial to analyze the effects of doxycycline in LID in patients with PD.

## MATERIALS AND METHODS

### Study design and participants

We performed an open-label, single-center, phase 2 proof-of-concept study to assess the efficacy and safety of doxycycline for 12 weeks in patients with PD and LID. Participants were recruited in the Movement Disorders Unit of Ribeirão Preto Medical School, Brazil, between October 2019 and May 2021. The study was conducted following the Declaration of Helsinki and Good Clinical Practice Guidelines and was approved by the Ribeirão Preto Medical School Ethics Committee (number 3.055.052). All patients provided written informed consent.

As inclusion criteria, we selected: patients aged 18 years or older; diagnosis of PD according to the UK Parkinson’s Disease Society Brain Bank clinical diagnostic criteria; at least a mild functional impact of dyskinesia in the Movement Disorder Society - Unified Parkinson’s Disease Rating Scale (MDS-UPDRS)^22^ Part IV (item 4.2 score > 1) at screening and baseline; use of levodopa at least three times daily; antiparkinsonian medications doses unchanged for at least four weeks before screening and during study participation.

As exclusion criteria, we excluded: atypical or secondary parkinsonisms; treatment of any experimental drug or intervention within 90 days before screening; moderate or severe psychotic symptoms (MDS-UPDRS Part I, item 1.2 score > 2); dementia according to MDS diagnostic criteria^23^; severe systemic conditions (infections, malignant neoplasms, chronic kidney or liver diseases); pregnancy or lactation; history of hypersensitivity or allergic reaction to tetracyclines; no clinical dyskinetic movements at baseline.

### Treatment

The study consisted of a 2-week titration phase and a 10-week maintenance phase. The dose of doxycycline 200 mg/day was chosen based on an optimized plasmatic concentration and usual dose in long-term treatments^24^. Initially, patients self-administrated one capsule of doxycycline 100 mg after breakfast once a day for two weeks; after two weeks of treatment, patients were evaluated by investigators in person; if investigators and patients reported no improvement in dyskinetic movements, the dose was increased to 100 mg b.i.d in week 3. After, patients were maintained on a constant dose of doxycycline until the end of week 12, when the drug was withdrawn. All antiparkinsonian medications were maintained unchanged until the end of the study.

### Assessments and outcome measures

Patients were evaluated at baseline and weeks 4 and 12 by a movement disorders specialist. Before the baseline visit, patients were trained in filling out the PD home diary, and diary concordance testing between patients and investigators was performed. Patients completed PD home diaries to assess their motor status every half hour for 24 hours for three consecutive days before baseline visit and before weeks 4 and 12 visits, and we calculated the three-day average from home diaries. Motor status was assessed as asleep, OFF time, ON time without dyskinesia, ON time with non-troublesome dyskinesia, and ON time with troublesome dyskinesia^25^. For analyses, the sum of ON time without dyskinesia with ON time with non-troublesome dyskinesia was called “ON time without troublesome dyskinesia.” At each visit, patients were instructed not to take their regular doses of antiparkinsonian drugs 12 hours before the evaluation and to be on an empty stomach.

As the first step of the baseline visit, patients were evaluated with the MDS-UPDRS in OFF state. After, patients ingested their regular dose of levodopa increased by 50% and were evaluated with the MDS-UPDRS Part III and the Unified Dyskinesia Rating Scale (UDysRS)^26^ in ON state; ON state evaluations were filmed for posterior assessment of two blinded investigators. Also, clinical and demographic data were recorded, and the completed PD home diary was reviewed. The same evaluations were performed at LID weeks 4 and 12, including the Clinical Global Impression of Change (CGIC) scale^27^ for patients and investigators regarding the change of intensity in dyskinesias compared to the baseline.

The primary outcome measure was the change from baseline in the UDysRS total score at week 12. Key secondary outcomes measures were OFF time and ON time with troublesome dyskinesia in the PD home diary. Other secondary outcomes measures included changes between baseline and week 12 visit in MDS-UPDRS Parts III and IV and total scores (ON state), ON time without troublesome dyskinesia in the PD home diary, and patient- and investigator-related CGIC.

### Statistical analysis

We performed a modified intention-to-treat analysis. Changes from baseline to weeks 4 and 12 in the outcomes (UDysRS, MDS-UPDRS, time measures from PD home diary) were assessed using the Friedman test to compare multiple repeated measures, and the Wilcoxon signed-rank test was used to compare baseline and week 12. Effect sizes were calculated based on the Wilcoxon signed-rank Z value by the square root of the number of related pairs. Missing data were imputed according to the last observation carried forward method.

## RESULTS

### Study population and baseline characteristics

A total of fifteen patients were screened, and eight patients met the inclusion criteria and were enrolled in the study (Figure 1). The most common reason for failure in enrollment was that patients (n = 5) had difficulties understanding the PD home diary filling procedures. Baseline demographics are provided in Table 1. The patients were predominantly women (6 women and two men), had long PD and LID duration (median 17 and 10 years, respectively), and used high doses of antiparkinsonian drugs (median 1162 mg/day). Four patients had moderate scores, and the other four had severe scores in time spent with dyskinesias on item 4.1 of the MDS-UDPRS. Two patients had mild scores, four had moderate scores, and the other two had severe scores in the functional impact of dyskinesias on item 4.2 of the MDS-UDPRS.

**Table 1.** Baseline clinical and epidemiological data of the eight patients.

|  |  |
| --- | --- |
| Male sex, % (n) | 25 (2) |
| Age at the time of evaluation (years) | 57.5 (53-65) |
| Age at onset of PD (years) | 41 (39-48) |
| Mini-Mental State Examination | 27 (25-29) |
| Disease duration (years) | 17 (8-21) |
| LID duration (years) | 10 (5-12) |
| Levodopa daily dose (mg/day) | 850 (724-1072) |
| Levodopa equivalent daily dose (mg/day) | 1162 (975-1701) |
| Concomitant medication use |  |
| Use of dopaminergic agonists, % (n) | 50 (4) |
| Use of amantadine, % (n) | 75 (6) |
| Use of MAO inhibitors, % (n) | 0 (0) |
| MDS-UPDRS |  |
| Part I | 11 (6-25) |
| Part II | 26 (15-35) |
| Part III (ON state) | 32 (27-38) |
| Part III (OFF state) | 53 (34-61) |
| Part IV | 10 (8-12) |
| Total score (ON state) | 77.5 (60-109) |
| Hoehn & Yahr (ON state) | 2 (2-3) |
| PD home diary |  |
| ON time with troublesome dyskinesia (hours) | 4.17 (2-7) |
| ON time without troublesome dyskinesia (hours) | 7.34 (4-10) |
| OFF time (hours) | 4.5 (1-5) |
| Participants with OFF time, % (n) | 100 (8) |
| UDysRS |  |
| Historical subscore | 31 (28-33) |
| Objective subscore | 36 (31-49) |
| Total score | 47.75 (46-59) |

Values are median (interquartile range) or % (n). Abbreviations: COMT, catechol-O-methyltransferase; LID, Levodopa-induced dyskinesias; MAO, monoamine oxidase; MDS-UPDRS, Movement Disorder Society–Unified Parkinson’s Disease Rating Scale; PD, Parkinson’s disease; UDysRS, Unified Dyskinesia Rating Scale.

**Figure 1.**
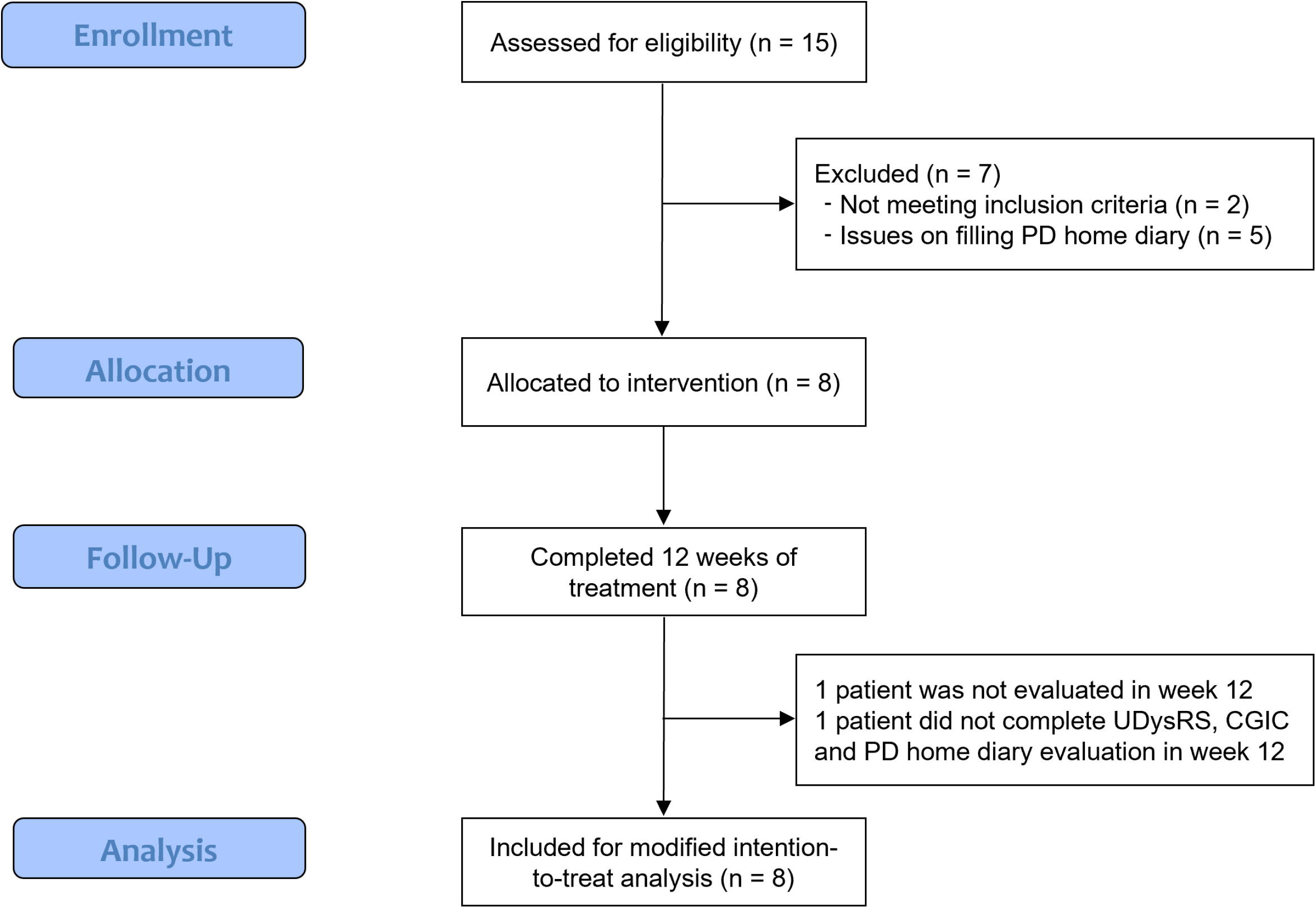
Fluxogram of the study. Abbreviations: CGIC, Clinical Global Impression of Change; PD, Parkinson’s disease; UDysRS, Unified Dyskinesia Rating Scale.

All patients received a daily dose of doxycycline 100 mg initially, and after two weeks, the daily dose was increased to 200 mg. All patients completed the treatment for 12 weeks, and all patients were evaluated at the baseline, at week 4 and week 12. One patient had issues reporting the PD home diary, and these data were excluded from the analysis.

### Outcomes

The primary efficacy analysis showed that the treatment with doxycycline was associated with a reduction in UDysRS total score until week 12 (Friedman’s X^2^ = 9.6, p = 0.008; week 12 compared to baseline, Wilcoxon Z = -2.521, p = 0.008; effect size = 0.89) (Figure 2, Table 2). There was a reduction in the historical subscore of the UDysRS until week 12 (Friedman’s X^2^ = 12.6, p = 0.002; week 12 compared to baseline, Wilcoxon Z = -2.524, p = 0.008), but not in the objective subscore (Friedman’s X^2^ = 1.31, p = 0.51).

**Table 2.**
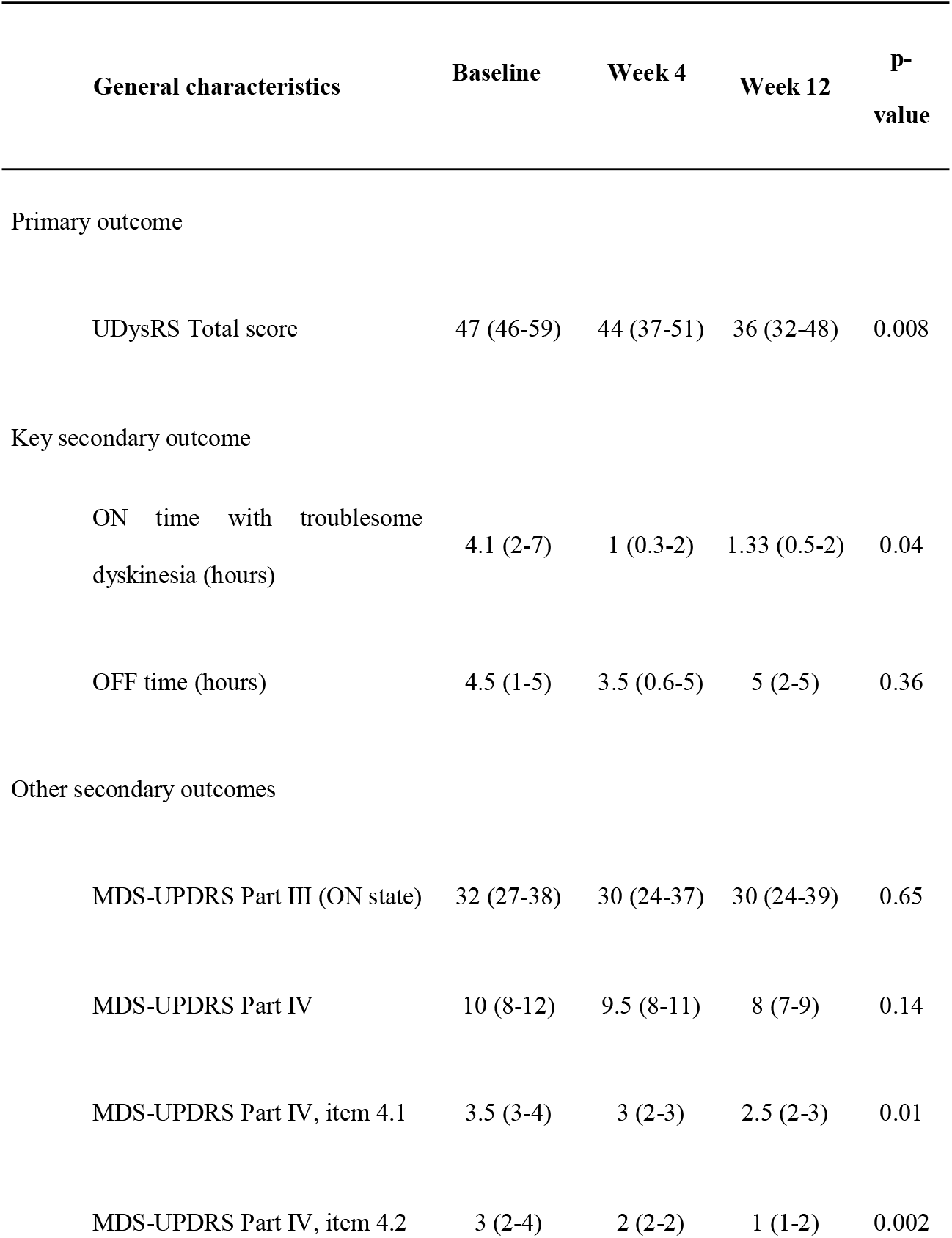
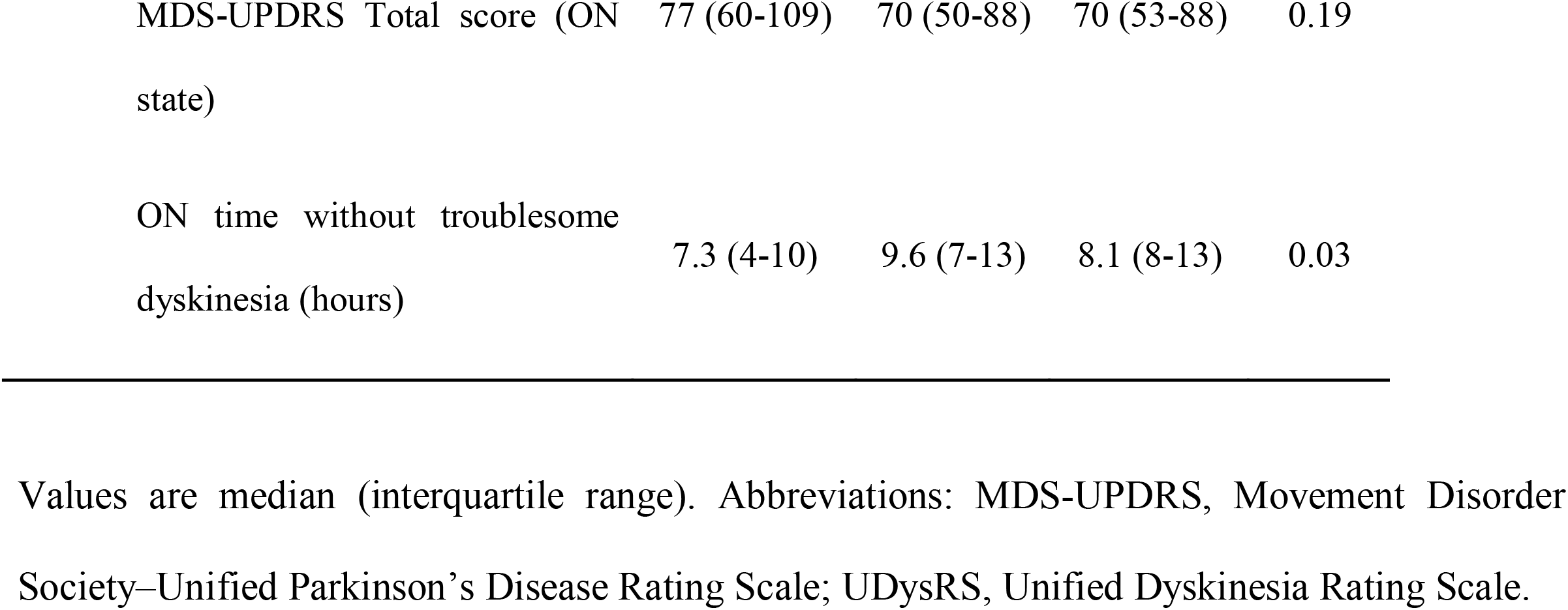
Efficacy results at the end of the treatment with doxycycline in the modified intention-to-treat population.

**Figure 2.**
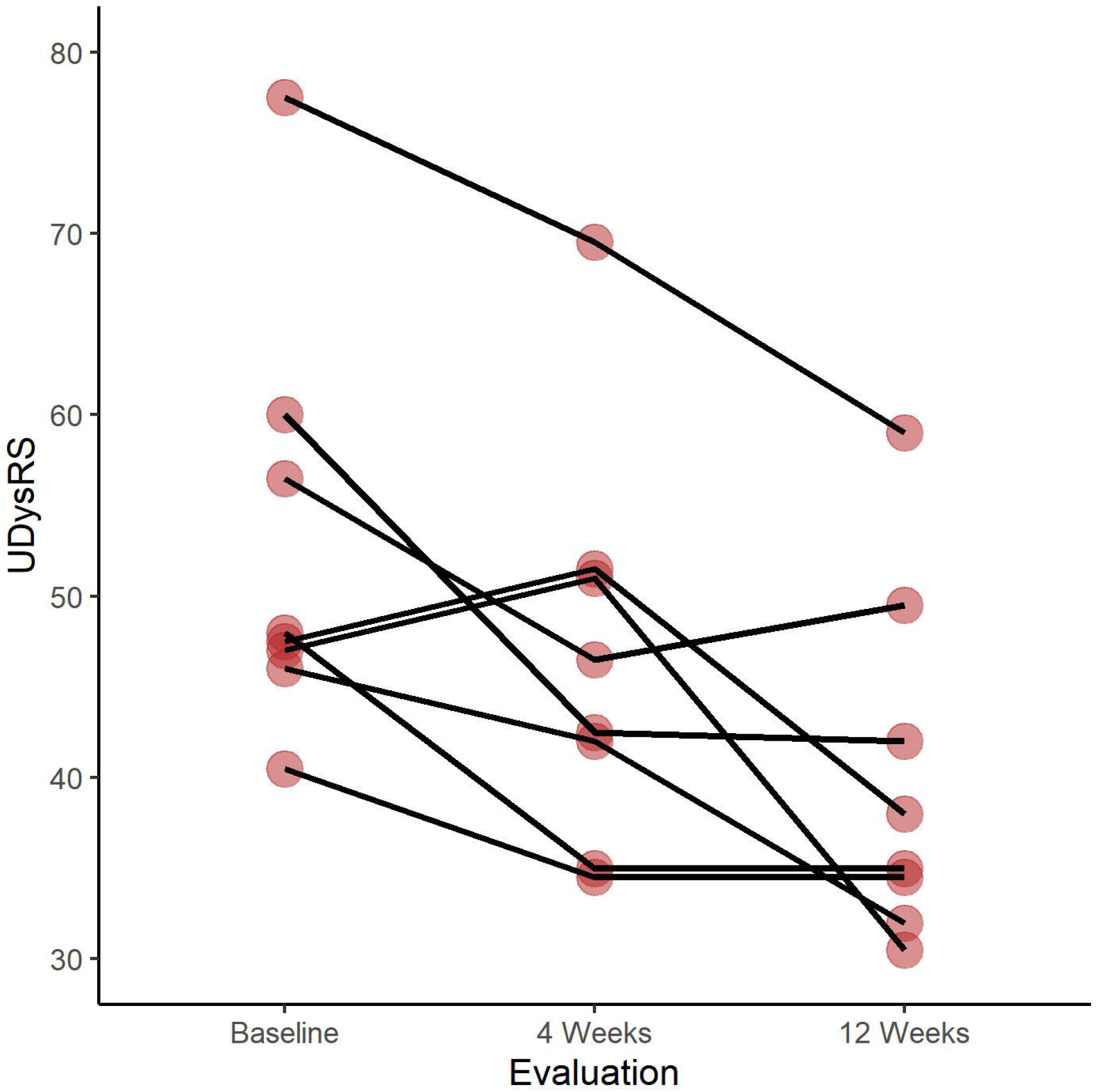
Change in the UDysRS total score over time (modified intention-to-treat population). Abbreviations: UDysRS, Unified Dyskinesia Rating Scale.

PD home diary measurements showed significant clinical improvements after treatment with doxycycline through 12 weeks: reduction of ON time with troublesome dyskinesias (Friedman’s X^2^ = 10.8, p = 0.004; week 12 compared to baseline, Wilcoxon Z = -2.36, p = 0.016; effect size = 0.89) and increase of ON time without troublesome dyskinesias (Friedman’s X^2^ = 6.74, p = 0.03; week 12 compared to baseline, Wilcoxon Z = -1.69, p = 0.1) (Figure 3, Table 2).

**Figure 3.**
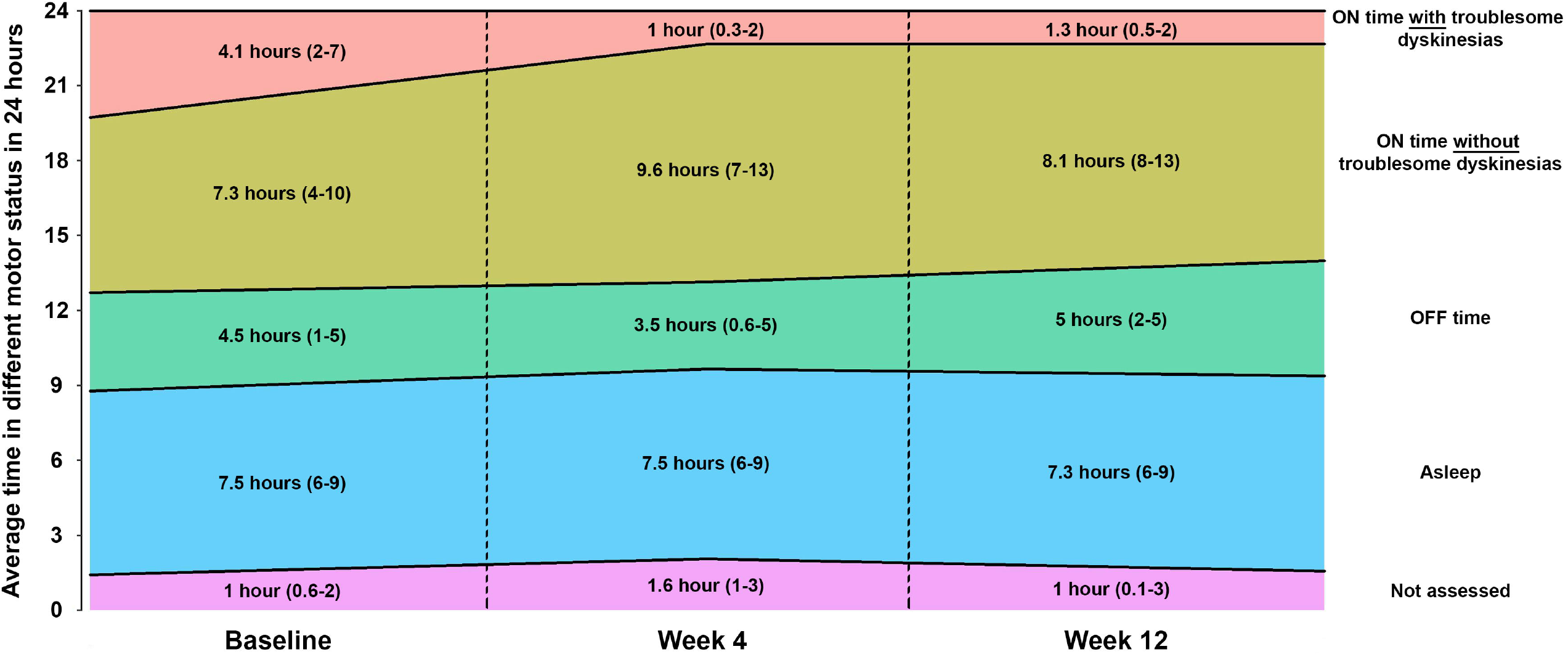
Change in the average time in different motor status according to the PD home diary over time (modified intention-to-treat population). Values are median (interquartile range).

There was no worsening of parkinsonism after 12 weeks of treatment with doxycycline measured by total score and Part III of the MDS-UPDRS in ON state (Table 2). There was no modification of the MDS-UPDRS Part IV after treatment, but there were significant reductions in scores of time spent with dyskinesias (item 4.1, Friedman’s X^2^ = 6.61, p = 0.03; week 12 compared to baseline, Wilcoxon Z = -2.07, p = 0.063) and functional impact of dyskinesias (item 4.2, Friedman’s X^2^ = 11.5, p = 0.003; week 12 compared to baseline, Wilcoxon Z = -2.56, p = 0.008).

The CGIC scale results showed an overall agreement between impressions from investigators and patients, with all patients assessed as improved by investigators after treatment with doxycycline at week 12, and seven of eight patients self-reported themselves as improved (87.5%).

### Safety

Overall, six patients (75%) reported at least one adverse event, one (12.5%) patient had a moderate adverse event, and no patient had an adverse event that led to discontinuation of the therapy or serious adverse events. Most adverse events were grade 1 or 2 according to the Common Terminology Criteria for Adverse Event (version 5.0). Most of these adverse events were drug-related, and dyspepsia (n = 4; all grade 1) and abdominal pain (n = 2; one grade 1 and one grade 2) were the most common events. Nausea (n = 1), headache (n = 1) and somnolence (n = 1) were also described.

## DISCUSSION

Doxycycline 200 mg daily for 12 weeks reduced LID frequency, severity, and functional impact in patients with PD using levodopa, as assessed by UDysRS, PD home diary, MDS-UPDRS, and CGIC. Also, doxycycline increased ON time without troublesome dyskinesias, without worsening motor and non-motor symptoms. From baseline, the UDysRS total score was 11 points lower and ON time without troublesome dyskinesias was 2.77 hours shorter in week 12. In the baseline, all patients with PD had LID during at least 50% of their waking day (MDS-UDPRS item 4.1 scores 3 and 4), and 75% of patients had LID, which impeded their activities (MDS-UDPRS item 4.2 scores 3 and 4); after doxycycline (week 12), only 50% of patients had LID during at least 50% of waking day, and no patient had LID which impeded activities.

The treatment with doxycycline 200 mg daily showed no serious adverse events. Dyspepsia, the most frequently observed adverse event, was mostly mild and did not cause discontinuation of the drug.

UDysRS total score has been suggested as the preferred primary outcome for clinical trials on LID^28^. Minimal clinically important difference (MCID) in LID has been explored recently. For UDysRS, there are established MCID for the historic subscores (Part 1 - ON dyskinesia, -2.1 points; Part 2 - OFF dyskinesia, -1.8 points)^29^ and the impairment subscore of the scale (Part 3, - 2.32 points)^30^. Our study showed a clinically important reduction of 10.5 points for the historic subscore Part 1 in week 12, but without significant reductions in Parts 2 and 3 of the UDysRS.

For comparison, the 274 mg extended-release oral formulation of amantadine (ADS-5102) once daily at bedtime was approved by the FDA in 2017 for the treatment of LID after positive results from two phase 3 randomized and controlled trials (EASE LID^31^ and EASE LID 3^32^). These studies showed a reduction of -15.9^31^ and -20.7 points^32^ in the UDysRS total score after ADS-5102 in week 12, as well as our doxycycline trial indicated a reduction of 11 points in the UDysRS in week 12. Also, ON time without troublesome dyskinesias increased in 3.6^31^ and 4 hours^32^ after ADS-5102 in week 12, and doxycycline increased the ON time without troublesome dyskinesias by 2.77 hours. A recent pooled analysis of these two studies calculated the magnitude of reduction of ON time with dyskinesias after ADS-5102 as a Cohen d effect size of 0.49^33^. For comparison, our effect size for reduction of ON time with troublesome dyskinesias after doxycycline was 0.83.

Another antibiotic has shown antidyskinetic properties: ceftriaxone slowed the development of abnormal involuntary movements but did not change previously established LID^34^, acting through the expression of glutamate transporter 1. Furthermore, doxycycline may have an additional positive effect in PD: inhibition of alpha-synuclein aggregation. Doxycycline inhibits the formation of toxic misfolded forms of alpha-synuclein oligomers through protein aggregation and blocks the seeding capacity of preformed aggregates in an in vitro and in vivo model^35^. Recently, another study in vivo confirmed these findings^36^.

A recent clinical trial showed that intestinal decontamination with colon enemas and the luminal antibiotic rifaximin reduced LID in patients with advanced PD^37^. The authors suggested that the therapy modified gut dysbiosis, common in PD^38^. Gut dysbiosis causes a systemic pro-inflammatory status that increases the brain-blood barrier permeability, promoting neuroinflammation together with bacterial products from the gut microbiota^39^. Doxycycline may also be effective in LID by reducing neuroinflammation mediated by gut dysbiosis.

As limitations, our study was not randomized and controlled, as well as the small size may have underpowered the results. Also, the short duration of treatment (12 weeks) may not be adequate to measure efficacy and safety for oral doxycycline 200 mg daily in LID. An additional group for doxycycline in a smaller daily dose could be included in further studies, considering that its antidyskinetic effect may not be associated with an antibiotic dose. The reduction of LID in rats after treatment with sub-antibiotic doses of doxycycline and with its derivative without antibiotic effect COL-3 might be used to reduce the risk of adverse events related to long-term therapy^21^.

In conclusion, this proof-of-concept clinical trial first revealed that doxycycline could directly modulate LID in accordance with previous preclinical studies. Therefore, further well-designed placebo-controlled clinical trials with a longer duration and a larger number of participants would be the next logical step in this field.

## Data Availability

All data produced in the present study are available upon reasonable request to the authors

## REFERENCES

1. GBD 2016 Parkinson’s Disease Collaborators. Global, regional, and national burden of Parkinson’s disease, 1990-2016: a systematic analysis for the Global Burden of Disease Study 2016. Lancet Neurol. 2018 Nov;17(11):939–53.

2. Bloem BR, Okun MS, Klein C. Parkinson’s disease. Lancet. 2021 Jun 12;397(10291):2284– 303.

3. Aquino CC, Fox SH. Clinical spectrum of levodopa-induced complications. Mov Disord. 2015 Jan;30(1):80–9.

4. Bortolanza M, Padovan-Neto FE, Cavalcanti-Kiwiatkoski R, Dos Santos-Pereira M, Mitkovski M, Raisman-Vozari R, et al. Are cyclooxygenase-2 and nitric oxide involved in the dyskinesia of Parkinson’s disease induced by L-DOPA? Philos Trans R Soc Lond B Biol Sci [Internet]. 2015 Jul 5;370(1672). Available from: http://dx.doi.org/10.1098/rstb.2014.0190

5. Mulas G, Espa E, Fenu S, Spiga S, Cossu G, Pillai E, et al. Differential induction of dyskinesia and neuroinflammation by pulsatile versus continuous l-DOPA delivery in the 6-OHDA model of Parkinson’s disease. Exp Neurol. 2016 Dec;286:83–92.

6. Julien C, Berthiaume L, Hadj-Tahar A, Rajput AH, Bédard PJ, Di Paolo T, et al. Postmortem brain fatty acid profile of levodopa-treated Parkinson disease patients and parkinsonian monkeys. Neurochem Int. 2006 Apr;48(5):404–14.

7. Santos-Lobato BL, Gardinassi LG, Bortolanza M, Peti APF, Pimentel ÂV, Faccioli LH, et al. Metabolic Profile in Plasma and CSF of levodopa-induced dyskinesia in Parkinson’s Disease: Focus on Neuroinflammation. Mol Neurobiol [Internet]. 2021 Dec 2; Available from: http://dx.doi.org/10.1007/s12035-021-02625-1

8. Santos-Lobato BL, Bortolanza M, Pinheiro LC, Batalhão ME, Pimentel ÂV, Capellari-Carnio E, et al. Levodopa-induced dyskinesias in Parkinson’s disease increase cerebrospinal fluid nitric oxide metabolites’ levels. J Neural Transm. 2022 Jan;129(1):55–63.

9. Del-Bel E, Bortolanza M, Dos-Santos-Pereira M, Bariotto K, Raisman-Vozari R. l-DOPA-induced dyskinesia in Parkinson’s disease: Are neuroinflammation and astrocytes key elements? Synapse. 2016 Dec;70(12):479–500.

10. Kong M, Ba M, Ren C, Yu L, Dong S, Yu G, et al. An updated meta-analysis of amantadine for treating dyskinesia in Parkinson’s disease. Oncotarget. 2017 Aug 22;8(34):57316–26.

11. Ossola B, Schendzielorz N, Chen S-H, Bird GS, Tuominen RK, Männistö PT, et al. Amantadine protects dopamine neurons by a dual action: reducing activation of microglia and inducing expression of GDNF in astroglia [corrected]. Neuropharmacology. 2011 Sep;61(4):574–82.

12. AlShimemeri S, Fox SH, Visanji NP. Emerging drugs for the treatment of L-DOPA-induced dyskinesia: an update. Expert Opin Emerg Drugs. 2020 Jun;25(2):131–44.

13. Johnston TH, Lacoste AMB, Visanji NP, Lang AE, Fox SH, Brotchie JM. Repurposing drugs to treat l-DOPA-induced dyskinesia in Parkinson’s disease. Neuropharmacology. 2019 Mar 15;147:11–27.

14. Bortolanza M, Santos-Lobato BL, Nascimento GC, Del-Bel E. Management with antibiotics in Parkinson’s disease [Internet]. Diagnosis and Management in Parkinson’s Disease. 2020. p. 491–510. Available from: http://dx.doi.org/10.1016/b978-0-12-815946-0.00029-6

15. Santa-Cecília FV, Socias B, Ouidja MO, Sepulveda-Diaz JE,Acuña L, Silva RL, et al. Doxycycline Suppresses Microglial Activation by Inhibiting the p38 MAPK and NF-kB Signaling Pathways. Neurotox Res. 2016 May;29(4):447–59.

16. Bortolanza M, Nascimento GC, Socias SB, Ploper D, Chehín RN, Raisman-Vozari R, et al. Tetracycline repurposing in neurodegeneration: focus on Parkinson’s disease. J Neural Transm. 2018 Oct;125(10):1403–15.

17. Egeberg A, Hansen PR, Gislason GH, Thyssen JP. Exploring the Association Between Rosacea and Parkinson Disease: A Danish Nationwide Cohort Study. JAMA Neurol. 2016 May 1;73(5):529–34.

18. Golub LM, Elburki MS, Walker C, Ryan M, Sorsa T, Tenenbaum H, et al. Non-antibacterial tetracycline formulations: host-modulators in the treatment of periodontitis and relevant systemic diseases. Int Dent J. 2016 Jun;66(3):127–35.

19. Lazzarini M, Martin S, Mitkovski M, Vozari RR, Stühmer W, Bel ED. Doxycycline restrains glia and confers neuroprotection in a 6-OHDA Parkinson model. Glia. 2013 Jul;61(7):1084–100.

20. Cox CA, Amaral J, Salloum R, Guedez L, Reid TW, Jaworski C, et al. Doxycycline’s effect on ocular angiogenesis: an in vivo analysis. Ophthalmology. 2010 Sep;117(9):1782–91.

21. Bortolanza M, do Nascimento GC, Raisman-Vozari R, Del-Bel E. Doxycycline and its derivative, COL-3, decrease dyskinesia induced by l-DOPA in hemiparkinsonian rats. Br J Pharmacol. 2021 Jul;178(13):2595–616.

22. Goetz CG, Tilley BC, Shaftman SR, Stebbins GT, Fahn S, Martinez-Martin P, et al. Movement Disorder Society-sponsored revision of the Unified Parkinson’s Disease Rating Scale (MDS-UPDRS): Scale presentation and clinimetric testing results [Internet]. Vol. 23, Movement Disorders. 2008. p. 2129–70. Available from: http://dx.doi.org/10.1002/mds.22340

23. Emre M, Aarsland D, Brown R, Burn DJ, Duyckaerts C, Mizuno Y, et al. Clinical diagnostic criteria for dementia associated with Parkinson’s disease. Mov Disord. 2007 Sep 15;22(12):1689–707; quiz 1837.

24. Beringer PM, Owens H, Nguyen A, Benitez D, Rao A, D’Argenio DZ. Pharmacokinetics of doxycycline in adults with cystic fibrosis. Antimicrob Agents Chemother. 2012 Jan;56(1):70–4.

25. Hauser RA, Friedlander J, Zesiewicz TA, Adler CH, Seeberger LC, O’Brien CF, et al. A home diary to assess functional status in patients with Parkinson’s disease with motor fluctuations and dyskinesia. Clin Neuropharmacol. 2000 Mar;23(2):75–81.

26. Goetz CG, Nutt JG, Stebbins GT. The Unified Dyskinesia Rating Scale: presentation and clinimetric profile. Mov Disord. 2008 Dec 15;23(16):2398–403.

27. Guy W. ECDEU assessment manual for psychopharmacology. 1976. 603 p.

28. Goetz CG, Stebbins GT, Chung KA, Hauser RA, Miyasaki JM, Nicholas AP, et al. Which dyskinesia scale best detects treatment response? Mov Disord. 2013 Mar;28(3):341–6.

29. Makkos A, Kovács M, Pintér D, Janszky J, Kovács N. Minimal clinically important difference for the historic parts of the Unified Dyskinesia Rating Scale. Parkinsonism Relat Disord. 2019 Jan;58:79–82.

30. Mestre TA, Beaulieu-Boire I, Aquino CC, Phielipp N, Poon YY, Lui JP, et al. What is a clinically important change in the Unified Dyskinesia Rating Scale in Parkinson’s disease? Parkinsonism Relat Disord. 2015 Nov;21(11):1349–54.

31. Pahwa R, Tanner CM, Hauser RA, Isaacson SH, Nausieda PA, Truong DD, et al. ADS-5102 (Amantadine) Extended-Release Capsules for Levodopa-Induced Dyskinesia in Parkinson Disease (EASE LID Study): A Randomized Clinical Trial. JAMA Neurol. 2017 Aug 1;74(8):941–9.

32. Oertel W, Eggert K, Pahwa R, Tanner CM, Hauser RA, Trenkwalder C, et al. Randomized, placebo-controlled trial of ADS-5102 (amantadine) extended-release capsules for levodopa-induced dyskinesia in Parkinson’s disease (EASE LID 3). Mov Disord. 2017 Dec;32(12):1701–9.

33. Hauser RA, Walsh RR, Pahwa R, Chernick D, Formella AE. Amantadine ER (Gocovri) Significantly Increases ON Time Without Any Dyskinesia: Pooled Analyses From Pivotal Trials in Parkinson’s Disease. Front Neurol. 2021 Mar 26;12:645706.

34. Chotibut T, Meadows S, Kasanga EA, McInnis T, Cantu MA, Bishop C, et al. Ceftriaxone reduces L-dopa-induced dyskinesia severity in 6-hydroxydopamine parkinson’s disease model. Mov Disord. 2017 Nov;32(11):1547–56.

35. González-Lizárraga F, Socías SB, Ávila CL, Torres-Bugeau CM, Barbosa LRS, Binolfi A, et al. Repurposing doxycycline for synucleinopathies: remodelling of α-synuclein oligomers towards non-toxic parallel beta-sheet structured species. Sci Rep. 2017 Feb 3;7:41755.

36. Dominguez-Meijide A, Parrales V, Vasili E, González-Lizárraga F, König A, Lázaro DF, et al. Doxycycline inhibits α-synuclein-associated pathologies in vitro and in vivo. Neurobiol Dis. 2021 Apr;151:105256.

37. Baizabal-Carvallo JF, Alonso-Juarez M, Fekete R. Intestinal Decontamination Therapy for Dyskinesia and Motor Fluctuations in Parkinson’s Disease. Front Neurol. 2021 Sep 10;12:729961.

38. Keshavarzian A, Green SJ, Engen PA, Voigt RM, Naqib A, Forsyth CB, et al. Colonic bacterial composition in Parkinson’s disease. Mov Disord. 2015 Sep;30(10):1351–60.

39. Baizabal-Carvallo JF, Alonso-Juarez M. The Link between Gut Dysbiosis and Neuroinflammation in Parkinson’s Disease. Neuroscience. 2020 Apr 15;432:160–73.

